# Zika Virus Infection: Review Of Neuroimage Studies And The Relationship Between Findings And Time Of Infection

**DOI:** 10.1101/19004283

**Authors:** Graciane Radaelli, Magda Lahorgue Nunes, Ricardo Bernardi Soder, Júlia Monteiro de Oliveira, Fernanda Thays Konat Bruzzo, Felipe Kalil Neto, Eduardo Leal Conceição, Mirna Wetters Portuguez, Jaderson Costa da Costa

## Abstract

**AIM:** To conduct a systematic literature review on neuroimage findings in children with microcephaly by Zika virus (ZIKV).

**METHOD:** We performed a literature search in PubMed, Cochrane Library and Web of Science for full-text articles reporting neuroimage exam of computed tomography scan or magnetic resonance imaging.

**RESULTS:** Were identified 2,214 publications. Of these 2,170 were excluded by the analysis of titles and abstracts, resulting in 7 articles included. The abnormalities presented in neuroimage showed the highest occurrence in the first trimester: decreased brain volume + increased extra-axial CSF space (100%), subcortical calcifications (89.1%), microcephaly (89.1%), ventriculomegaly (72.9%), malformation of cortical development (40.5%), basal ganglia calcifications (40.5%), megacisterna magna (39.1%). In relation to the second trimester of ZIKV infection, the most common were as follows: decreased brain volume + increased extra-axial CSF space (100%), subcortical calcifications (100%), microcephaly (85.7%), ventriculomegaly (71.4%), malformation of cortical development (71.4%), basal ganglia calcifications (19%), megacisterna magna (4.7%). In relation to the neuroimage abnormalities detected in the 3rd trimester 2 cases were found.

**INTERPRETATION:** This systematic review is the first that evaluates brain changes in newborns with different neuroimage techniques (CT and MRI exams) and related findings with the gestational period of ZIKV infection.

## INTRODUCTION

Many original articles and case series have been published emphasizing neuroimage findings on congenital ZIKV infection. The overwhelming majority of these studies do not follow a neuroradiological methodology to describe the malformations and brain abnormalities resulting from ZIKV infection, much less a cause-and-effect correlation between the gestational period of maternal infection and the severity of encephalic changes at birth. Neuroimage methods with different sensitivities and diagnostic accuracy have been employed at random to elaborate tables and descriptive results of the main neuroradiological changes present in the newborns with congenital ZIKV infection. The consequence of this is the lack of a specific neuroradiological signature that relates to the gestational trimester of fetal infection by ZIKV.

ZIKV is a flavivirus transmitted mainly by mosquitoes of the genus Aedes. The virus was first isolated in Uganda in 1947 and remained restricted to areas in Africa and Asia for sixty years. In early 2015, in Brazil, however, there was an outbreak of ZIKV infection in the northeast region of the country [1]. In October 2015, an increase in the number of cases of microcephaly was reported, and this congenital malformation was related to intrauterine ZIKV infection [2,3]. The most frequent symptoms of ZIKV are fever, myalgia and rash [4]. ZIKV is usually diagnosed by the set of symptoms and standard reverse transcriptase polymerase chain reaction (RT-PCR) from blood, urine or saliva fluids. However, there are some difficulties in the diagnosis due to possible cross-reaction with other flaviviruses [5]. In relation to neuroimage findings, a variety of cortical malformations can be identified. They include parenchymal volume reduction, alterations in the gyral pattern, as well in white and gray matter, ventriculomegaly, abnormalities of the corpus callosum and cerebellum, hypoplasia and / or atrophy of the brainstem and difuse calcifications [6]. The World Health Organization (WHO) reported in 2016 cases of microcephaly and neurological disorders associated to ZIKV as a public health emergency of international concern, intensifying the need for further research in the area [7]. Microcephaly may not be evident at birth but may develop after birth along with other brain abnormalities. Therefore, it is essential that infants exposed to the ZIKV do perform neuroimage exams investigation. The objective of the present study was to conduct a systematic review of the literature to evaluate radiological findings and to create a neuroimage profile of children with microcephaly exposed to intrauterine ZIKV infection regarding the time of intrauterine infection.

## METHOD

This systematic review was performed following the methodology described in the Cochrane Handbook for Systematic Reviewers and presented by Preferred Reporting Items for Systematic Review and Meta-analyzes: the PRISMA Statement [8]. The search protocol was designed and registered in PROSPERO (CRD42017058801).

### Search strategy

We performed a literature search in PubMed, Cochrane Library and Web of Science. Following terms were used in the search strategy: “Zika virus”, “microcephaly”, “newborn”, “pregnancy” and “neuroimage”.

### Eligibility criteria

Inclusion criteria: we included prospective and retrospective clinical studies with neuroimage exams: computed tomography (CT) scan or magnetic resonance imaging (MRI) of individuals infected with ZIKV with microcephaly without language or year restriction until march 2018.

Exclusion criteria: Preclinical studies, in vivo studies, letters to the editor, editorials, duplicate publications, articles that do not have data to be evaluated, or articles that do not address neuroimage (CT or MRI) were excluded.

### Data extraction

The databases were searched and duplicate entries were removed. Abstracts that did not provide sufficient information regarding the inclusion and exclusion criteria were selected for full-text evaluation. In the second phase, the same reviewers independently evaluated the full text of these articles and made their selection in accordance with the eligibility criteria. Two reviewers (JM and BBF) performed the literature search and study selection independently. Disagreements were solved by consensus or by a third reviewer (GR).

#### Statistical analysis

To verify the statistically significant differences of the neuroradiological findings between the cases of ZIKV infection in the first and second trimester of gestation, we performed a Chi-squared-exact fisher test with a 95% confidence interval.

## RESULTS

The initial database resulted in 2,214 articles. After the removal of duplicate files, 2,211 articles were filtered according to the inclusion criteria. Of these 2,170 were excluded by the analysis of titles and abstracts, resulting in 41 articles for evaluation of the full text. 17 documents were excluded because there was no investigation with CT or MRI and 2 because they were experimental studies, resulting in a total of 22 studies. Subsequently, 11 articles were excluded because they were not a case report, and 4 were excluded because they did not include the trimester of gestation that occurred the infection, totalizing 7 articles included [9-15]. The complete flowchart is shown in Figure 1.

**Fig. 1.**
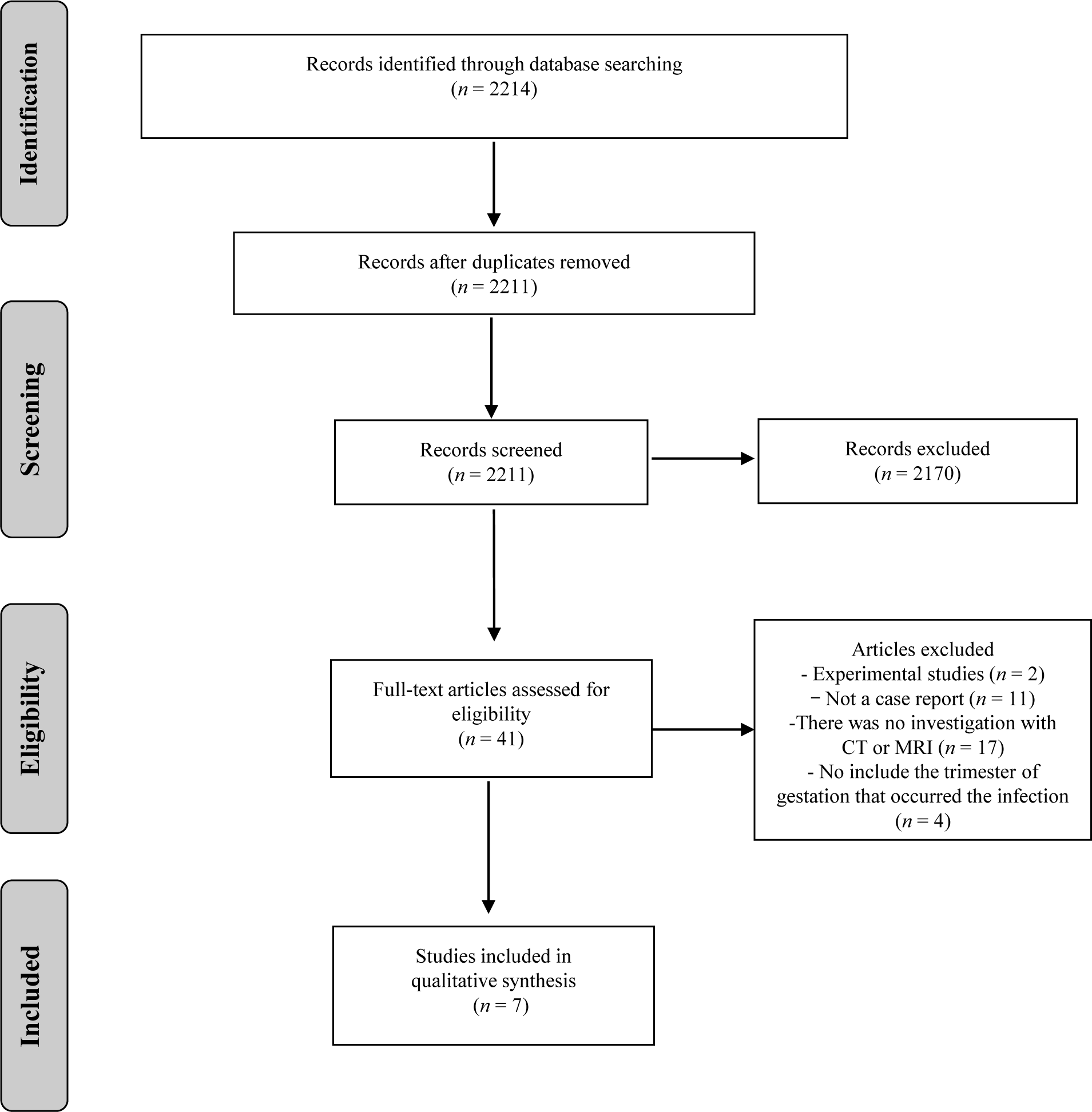
Summary of evidence search and study selection.

Our review included 125 cases of congenital Zika syndrome (CZS) reported in 7 studies (Table 1). There was a predominance of cases described in the first trimester (n=74) when compared to the second (n=21) and the third (n=2). There was a predominance of neuroimage abnormalities in the first and second trimester as described in Table 2 and Fig 2. The most frequent findings in the first and second trimester (1^st^ and 2^nd^) were decreased brain volume + increased extra-axial CSF space (100% and 100%), subcortical calcifications (89.1% and 100%), microcephaly (89.1% and 85.7), ventriculomegaly (72.9% and 71.4%)), malformation of cortical development (40.5% and 71,4%), basal ganglia calcifications (40.5% and 19%), megacisterna magna (39.1% and 4.7%). In the 3rd trimester, the first case, the mother was infected in the 7th month of gestation, presenting cortical calcifications with cortical volumetric reduction and in the second case, the infection occurred in the 8th month of gestation and neuroimage showed subcortical calcifications, reduced cortical volume, and colpocephaly.

**Table 1.** Summary of the time of infection with Congenital Zika Syndrome.

| Author(s), year | No. of cases<br>(*CT, MR or both) | Time of Infection (maternal symptoms) |  |  |  |
| --- | --- | --- | --- | --- | --- |
|  |  | 1° Trimester | 2° Trimester | 3° Trimester | Unknown |
| *Aragao et al. (2016)* | 23<br>(15, 1, 7) | 17 | 5 | --- | 1 |
| *van der Linden, (2016)* | 13<br>(3, 0, 10) | 4 | 2 | --- | 7 |
| *Aragao et al. (2017)* | 19<br>(0, 19, 0) | 9 | 5 | --- | 5 |
| *Carvalho et al. (2017)* | 37<br>** | 18 | 4 | 2 | 13 |
| *Castro et al. (2017)* | 8<br>(0, 7, 0) | 7 | --- | --- | 1 |
| *Pires et al. (2017)* | 8<br>(8, 0, 0) | 5 | 2 | --- | 1 |
| *Parra-Saavedra et al. (2017)* | 17<br>(0, 16, 0) | 14 | 3 | --- | --- |
| **Total of cases** | 125 | 74 | 21 | 2 | 28 |

\*CT = computed tomography; MRI = magnetic resonance imaging; \*\* The study does not specify RM or CT or both;

**Table 2.** Neuroimage findings: First and Second Trimester.

| Neuroimage findings | Patient infected in First Trimester<br>N=74 | Patients infected in Second Trimester<br>N=21 | OR (IC95%) | Chi-squared test Exacto de Fisher<br>(*p*) |
| --- | --- | --- | --- | --- |
| *Decreased brain volume + Increased extra-axial CSF space* | 74 (100) | 21 (100) | - | - |
| *Subcortical calcifications* | 66 (89.1) | 21 (100) | - | 0.248 |
| *Microcephaly* | 66 (89.1) | 18 (85.7) | 1,37 (0.33 -5.75) | 0.912 |
| *Ventriculomegaly* | 54 (72.9) | 15 (71.4) | 1.08 (0.36 – 3.17) | 0.999 |
| *Malformation of cortical development* | 30 (40.5) | 15 (71.4) | 0.27 (0.09 – 0.78) | **0.02\*** |
| *Basal ganglia calcifications* | 30 (40.5) | 4 (19) | 2.89 (0.88 – 9.46) | 0.113 |
| *Megacisterna magna* | 29 (39.1) | 1 (4.7) | 12.89 (1.64 – 101.3) | **0.002\*** |
| **Corpus callosum abnormalities** | 28 (37.8) | 8 (38) | 0.98 (0.36 – 2.68) | 0.999 |
| **Brainstem hypoplasia** | 22 (29.7) | 2 (9.5) | 4.0 (0.86 – 18.75) | 0.097 |
| **Periventricular calcifications** | 21 (28.3) | 3 (14.2) | 2.37 (0.63 – 8.92) | 0.303 |
| **Cerebellum hypoplasia/atrophy** | 19 (25.6) | 3 (14.2) | 2.07 (0.54 – 7.82) | 0.431 |
| **Brainstem calcification** | 12 (16.2) | 1 (4.7) | 3.87 (0.47 – 31.65) | 0.324 |
| **Cerebellum calcification** | 3 (4) | 3 (14.2) | 0.25 (0.04 – 1.36) | 0.239 |

*Data are presented as counts (percentage)*

**Fig 2.**
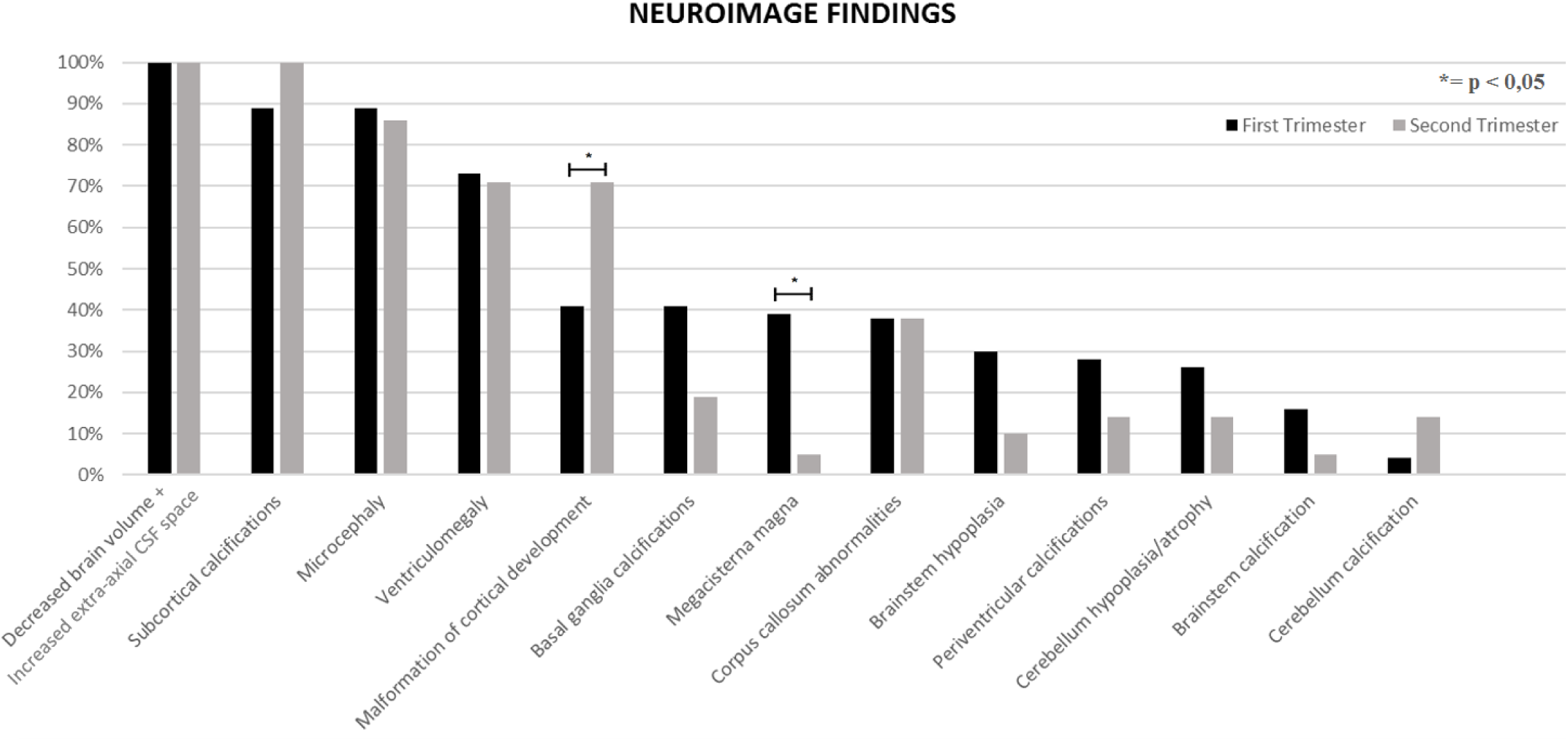
Neuroimage findings: first and second trimester.

Two outcomes had a score of p <0.05: Megacisterna magna (p = 0.002), showing a more frequent symptom when contamination occurred in the first trimester and Malformation of cortical development (p = 0.02), which was more recurrent in cases of contamination in the second trimestrer (Table 2).

## DISCUSSION

This systematic review is the first that evaluates brain changes in newborns with congenital infection by ZIKV with distinct neuroimage methods (CT and MRI exams) and treied to associated abnormalities with the gestational period of infection.

The studies published so far has described image abnormalities at random, without pre-established neuroradiological criteria, using imaging modalities with different sensitivity and accuracy, which compromise a reliable and adequate statistical analysis. Another limitation of the great majority of the published studies concerns the absence of the cause-effect relationship, due to the non-description of the gestational period that the fetal infection occurred. In addition, the present systematic review found numerous neuroimage findings only describes numerous cerebral malformations with microcephaly and at no time evidence the severity of the outcomes with the period of ZIKV infection. The main finding of this systematic review was that this review showed that the most serious and prevalent alterations occurred when the infection occurred in the first trimester of gestation. In contrast, it was verified that infections occurred in the second and third trimester of gestation presented less severe and complex radiological alterations (table 1).

Thus, because ZIKV interferes with the process of neuronal migration and consequent brain formation, the most prevalent radiological alterations, both in the first and second trimester, were subcortical calcifications, ventriculomegaly, microcephaly, cerebral volume decrease and cortical development malformation. Regarding patients with infections in the third trimester, only two cases of microcephaly were described and no additional malformations were reported.

The possible relationship of ZIKV infection during pregnancy and the aforementioned CNS changes gives rise to great concern for global public health, as this virus presently does not have any effective treatment. The period of infection of the ZIKV and the occurrence of fetal brain abnormalities that may be susceptible to viral pathogenesis, and how it affects the appearance of brain abnormalities on imaging, are currently poorly understood.

Brain changes caused by infectious agents during pregnancy differ from those that occur in children and adults because they occur during nervous tissue development. Manifestations of these infections will differ depending on the time when they occur and are less related to the virulence of the agent. Thus, in general, infections during the first two trimesters will result in congenital malformations, while those occurring in the third trimester will result in destructive lesions [16].

The ZIKV infection causes abnormalities during neural differentiation. Neural stem cells (NSCs) represent a primary target for ZIKV infection. During the infection, ZIKV proteins interfere in the NSC function causing severe consequences such as metabolic fluxes, inhibition of cell proliferation, and cellular apoptosis [17]. Neurospheres generated from human Induced Pluripotent Stem Cells (iPSC) after 3 days of ZIKV infection showed a morphological alteration and cell detachment, which can be associated mainly with cellular migration processes. After 6 days of exposure to ZIKV, only few neurospheres survived to the virus action. ZIKV can be observed in membranes, mitochondria and vesicles of cells. Other alterations caused by ZIKV in neurospheres include apoptotic nuclei formation, hallmark of cell death and presence of smooth membrane. The impact of ZIKV during embryonic neurogenesis was showed by the exposition of ZIKV to cerebral organoids derived from iPSC. The average growth area of the organoids exposed to the ZIKV was reduced by 40% when compared to the control cerebral organoids. This result demonstrated that ZIKV induces cell death in human iPS-derived neural stem cells, disrupts the formation of neurospheres and reduces organoids growth, indicating that ZIKV infection in models that mimics the first trimester of brain development may result in severe damage [18].

Similar studies showed that ZIKV infection can modulate cellular metabolism in neuronal cells in the first stages of neurodifferentiation through P53 activation and mTOR pathway inhibition by induction of early shifting from glycolysis to OXPHOS that produce immature differentiation, apoptosis, and stem cell exhaustion [19, 20].

Another important contribution of the current systematic review, which may be of great value to subsequent studies, is the discovery of numerous methodological or descriptive limitations that may have distorted the prevalence or frequency of neuroradiological findings in newborns with congenital ZIKV. One of the most important limitations was the absence of neuroradiological criteria for the description of the brain abnormalities, which can be verified by the absence of homogeneity of the findings, often using two descriptions for the same neuroradiological finding. Another relevant limitation, which certainly modified the actual frequency / prevalence of described neuroradiological abnormalities, was the equivalence between ultrasonography (US), CT and MRI during the description and tabulation of the results, without taking into account the disparity in sensitivity and accuracy between the three methods of which resulted in high frequencies and a greater diversity of brain changes in studies with higher number of MRIs and few changes / descriptions in those studies with few MRI exams. The difficulties encountered in assessing the relationship between the gestational age at which ZIKV infection occurred and the structural changes in the brain are associated with a lack of laboratory evidence and recorded clinical information, imaging studies of different sensitivities and/or reports with limited detail of the abnormalities and the heterogeneity in the their description in some cases. Also, in some articles in addition, in some reports there is no description of changes in the midline abnormalities such as changes in the corpus callosum and cerebellar vermis (perhaps due to the predominant number of CT scans and low quality MRI) and no recognition of cerebral and cerebellar atrophy or enlargement of cerebrospinal fluid space. Lastly, poor experience of the radiologist with pediatric neuroimage, many of which were interpreted by non-radiologists or general radiologists.

The number of studies available in the literature specifying radiological findings of congenital ZIKV infection still scarce. The clinical and radiological peculiarities of this pathology are in constant debate, as our review demonstrates. The necessity for a final description and classification of ZIKV brain abnormalities, using appropriate neuroimage criteria that allow a temporal cause-effect relationship with the gestational period of fetal infection may be of great value in assessing the outcomes and prognoses of affected children. Magnetic resonance imaging (MRI) is the best method for assessing the central nervous system (CNS) of fetuses with suspected defects. The absence of ionizing radiation and the ability for greater tissue differentiation allows for a better study of the CNS, without the limitations caused by skull artifact or fetal position. Further MRI studies are required for a more detailed and specific evaluation of the brain abnormalities caused by the congenital ZIKV infection, studies that comprise a more homogenous design, with a higher number of evaluated outcomes and that have two independent evaluators and trained to perform the image exams.

## CONCLUSION

Neuroimage abnormalities are much more prevalent and severe when the infection occurred in the 1^st^, 2^nd^ trimester in comparison to the 3^rd^. Findings that might characterized /differentiate infections that occurred in the first trimester in comparison to the second are malformation of cortical development and megacisterna magna.

## Data Availability

Not Applicable.

## Data Availability

Not Applicable.

## CONFLICT OF INTEREST

The authors have no conflicts of interest to disclose.

## ETHICAL PUBLICATION STATEMENT

We confirm that we have read the Journal position on issues involved in ethical publication and affirm that this report is consistent with those guidelines.

## ACKNOWLEDGMENTS

JCC and MLN are funded by CNPq (bolsa de produtividade em pesquisa). JCC and MLN are researchers respectively 1A and 1D from CNPq - Brazil. Our study was supported by the following grants: CNPq (Conselho Nacional de Desenvolvimento Científico e Tecnológico); Coordenação de Aperfeiçoamento de Pessoal de Nível Superior (CAPES). We thank Daniel Rodrigo Marinowic, PhD for his suggestion regarding in vitro studies.

## FUNDING SOURCES

This research did not receive any other specific grant from funding agencies in the public, commercial, or not-for-profit sectors.

## Notes

### Competing Interest Statement

The authors have declared no competing interest.

### Author Declarations

All relevant ethical guidelines have been followed and any necessary IRB and/or ethics committee approvals have been obtained.

Any clinical trials involved have been registered with an ICMJE-approved registry such as ClinicalTrials.gov and the trial ID is included in the manuscript.

## REFERENCES

(1) Desai, S. K., Hartman, S. D., Jayarajan, S., Liu, S., & Gallicano, G. I. (2017). Zika Virus (ZIKV): a review of proposed mechanisms of transmission and associated congenital abnormalities. American journal of stem cells, 6(2), 13.

(2) Mehrjardi, M. Z., Carteaux, G., Poretti, A., Taheri, M. S., Bermudez, S., Werner, H., & da Cruz Jr, L. C. H. (2017). Neuroimage findings of postnatally acquired Zika virus infection: a pictorial essay. Japanese journal of radiology, 35(7), 341–349.

(3) Nunes M.L., Carlini C.R., Marinowic D., Neto F.K., Fiori H.H., Scotta M.C., Zanella P.L., Soder R.B., da Costa J.C. Microcephaly and Zika virus: a clinical and epidemiological analysis of the current outbreak in Brazil. J Pediatr (Rio J). 2016.

(4) de Oliveira, W. K., Carmo, E. H., Henriques, C. M., Coelho, G., Vazquez, E., Cortez-Escalante, J., … & Dye, C. (2017). Zika virus infection and associated neurologic disorders in Brazil. New England Journal of Medicine, 376(16), 1591–1593.

(5) Paiva, M. H. S., Guedes, D. R. D., Leal, W. S., & Ayres, C. F. J. (2017). Sensitivity of RT-PCR method in samples shown to be positive for Zika virus by RT-qPCR in vector competence studies. Genetics and molecular biology, (AHEAD), 0-0.

(6) de Oliveria Szejnfeld, P. S., Levine, D., & de Oliveira Melo, A. S. (2016). Congenital brain abnormalities and Zika virus: what the radiologist can expect to see prenatally and postnatally. Radiology, 281, 1–16.

(7) de Oliveira, W. K., de França, G. V. A., Carmo, E. H., Duncan, B. B., de Souza Kuchenbecker, R., & Schmidt, M. I. (2017). Infection-related microcephaly after the 2015 and 2016 Zika virus outbreaks in Brazil: a surveillance-based analysis. The Lancet, 390(10097), 861–870.

(8) Moher, D., Liberati, A., Tetzlaff, J., Altman, D. G., & Prisma Group. (2009). Preferred reporting items for systematic reviews and meta-analyses: the PRISMA statement. PLoS medicine, 6(7), e1000097.

(9) Aragao, M. D. F. V., van der Linden, V., Brainer-Lima, A. M., Coeli, R. R., Rocha, M. A., da Silva, P. S., … & Valenca, M. M. (2016). Clinical features and neuroimage (CT and MRI) findings in presumed Zika virus related congenital infection and microcephaly: retrospective case series study. BMJ, 353, i1901.

(10) van der Linden, V. (2016). Description of 13 infants born during October 2015– January 2016 with congenital Zika virus infection without microcephaly at birth— Brazil. MMWR. Morbidity and mortality weekly report, 65.

(11) Aragao, M. F. V. V., Holanda, A. C., Brainer-Lima, A. M., Petribu, N. C. L., Castillo, M., van der Linden, V., … & Sarteschi, C. (2017). Nonmicrocephalic infants with congenital Zika syndrome suspected only after neuroimage evaluation compared with those with microcephaly at birth and postnatally: how large is the Zika virus “iceberg”?. American Journal of Neuroradiology, 38(7), 1427–1434.

(12) Carvalho, M. D. C. G., de Barros Miranda-Filho, D., van der Linden, V., Sobral, P. F., Ramos, R. C. F., Rocha, M. Â. W., … & Nunes, M. L. (2017). Sleep EEG patterns in infants with congenital Zika virus syndrome. Clinical Neurophysiology, 128(1), 204–214.

(13) Castro, J. D. V. D., Pereira, L. P., Dias, D. A., Aguiar, L. B., Maia, J. C. N., Costa, J.I F. D., … & Carvalho, F. H. C. (2017). Presumed Zika virus-related congenital brain malformations: the spectrum of CT and MRI findings in fetuses and newborns. Arquivos de Neuro-Psiquiatria, 75(10), 703–710.

(14) Pires, P., Jungmann, P., Galvão, J. M., Hazin, A., Menezes, L., Ximenes, R., … & Júnior, E. A. (2018). Neuroimage findings associated with congenital Zika virus syndrome: case series at the time of first epidemic outbreak in Pernambuco State, Brazil. Child’s Nervous System, 34(5), 957–963.

(15) Parra-Saavedra, M., Reefhuis, J., Piraquive, J. P., Gilboa, S. M., Badell, M. L., Moore, C. A., … & Sanz-Cortes, M. (2017). Serial head and brain imaging of 17 fetuses with confirmed Zika virus infection in Colombia, South America. Obstetrics & Gynecology, 130(1), 207–212.

(16) Bale JF Jr. Fetal infections and brain development. ClinPerinatol. 2009;36(3):639–53. https://doi.org/10.1016/j.clp.2009.06.005.

(17) Rothan H.A., Fang S., Mahesh M., Byrareddy S.N. Zika Virus and the Metabolism of Neuronal Cells. Molecular Neurobiology. doi.org/10.1007/s12035-018-1263-x

(18) Garcez P. P., Loiola E. C., da Costa R. M., Higa L. M., Trindade P., Delvecchio R., Nascimento J. M., Brindeiro R., Tanuri A., Rehen S. K. Zika virus impairs growth in human neurospheres and brain organoids. Science. 10.1126/science.aaf6116 (2016).

(19) Ghouzzi VE, Bianchi FT, Molineris I, Mounce BC, Berto GE, Rak M, Lebon S, Aubry L et al. ZIKA virus elicits P53 activation and genotoxic stress in human neural progenitors similar to mutations involved in severe forms of genetic microcephaly and p53. Cell Death Dis 7:e2440.

(20) Liang Q, Luo Z, Zeng J, Chen W, Foo SS, Lee SA, Ge J,Wang S et al. Zika virus NS4A and NS4B proteins deregulate Aktm TOR signaling in human fetal neural stem cells to inhibit neurogenesis and induce autophagy. Cell Stem Cell 19:663–671.

